# User-Informed Design of a Psychological Support Social Pet Robot for Community-Dwelling People With Long-Term Physical Disabilities: A Formative Study

**DOI:** 10.64898/2026.11.09.26367593

**Authors:** Yufei Lu, Xiaobin Chen, Ruomu Hui, Yushen Dai, Xiaoxiao Mei, Ho So, Chen Li, Xiaoming Tao, Yan Li

## Abstract

**Background:** Community-dwelling people with long-term physical disabilities may experience pain that interferes with daily and social activities and is associated with poorer mental wellbeing. A home-based psychological support social pet robot could offer mood check-ins, companionship, supportive interaction, and mindfulness guidance for community-dwelling people with long-term physical disabilities. Preliminary evidence is needed to understand their preferences and translate them into candidate design requirements.

**Objective:** This study aimed to identify preliminary needs and preferences to inform a psychological support social pet robot for community-dwelling people with long-term physical disabilities, offering mood check-ins, companionship, and mindfulness guidance at home.

**Methods:** We conducted a 3-phase formative user-informed design study comprising initial concept development, qualitative target user interviews, and a design workshop. Six community-dwelling adults with long-term physical disabilities took part in the interviews, and one participant also answered the workshop questions by telephone. Four authors used modified workshop materials to synthesize the interview findings and separate responses into an early product plan.

**Results:** Qualitative content analysis of the interview transcripts produced 5 themes. These were emotional support, soft touch and appearance, mindfulness delivery, context of use, and ease of use and accessibility. Participants expressed preferences about companionship, touch, use settings, and interaction effort. The author workshop combined these findings with author and technical judgments to develop candidate requirements for dialogue and mindfulness delivery. The resulting psychological support concept combined mood check-ins and companion-oriented interaction with flexible mindfulness guidance. The design also needed to consider pain and daily routines. After the workshop, the authors developed a physical proof-of-concept prototype. It implemented only part of the candidate requirements and included additional author or technical choices.

**Conclusions:** For this small community-dwelling sample, target user input showed that home use, physical access, companionship, and flexible support were important. Mindfulness was one component of the broader psychological support role, alongside mood check-ins and optional companion interaction. The prototype illustrates but does not validate the early requirements. Further studies should include people with a wider range of long-term physical disabilities and test usability, accessibility, privacy, and nursing value.

## Introduction

### Background

Community-dwelling people with long-term physical disabilities may experience pain and functional limitations alongside difficulties with mental wellbeing and social participation [1,2]. A study of adults with spinal cord injury or multiple sclerosis and chronic pain found that greater pain was associated with more interference in daily and social activities and poorer psychological functioning [1]. A longitudinal study of adults with four long-term disabling conditions found that functional impairment and problems related to mood, fatigue, and sleep were associated with lower satisfaction with social roles [2]. Accessible solutions are needed to support mental wellbeing in daily life. One clinical study linked pain to limits in daily activities and social participation [3]. Loneliness is linked to depression and anxiety in adults [4,5]. Another clinical study linked depression and anxiety to treatment response and later physical health [6].

The intended design aims to provide accessible psychological support at home alongside professional care. Self-management recommendations for a specific clinical group address physical and psychological needs [7]. Mindfulness is one possible form of support. It involves paying attention to current experience with an open and nonjudgmental attitude [8]. Mindfulness interventions have been studied as ways to support stress management and emotional wellbeing in a specific clinical group [9,10]. In this study, mindfulness was considered one component of a broader psychological support role that also included mood check-ins and companionship. Nursing recommendations for a specific clinical group identify psychosocial support, self-management, and needs-based patient education as parts of care [11,12]. These clinical recommendations help explain possible links between home support and professional care.

Social robots are physical systems that interact with people in social ways [13,14]. They can communicate through voice, movement, visual displays, and touch [14]. One set of clinical self-management solutions includes digital health tools, such as mobile applications where appropriate and available [7]. Chatbots can also support text or voice conversations. However, chatbots usually work through a phone or computer screen and do not have a physical body in the user environment [15]. A social robot combines conversation with physical form and social cues. Social pet robot refers to an animal-shaped social robot intended for companion-oriented interaction. Beyond conversation through applications, chatbots, or smart speakers, a soft, touchable animal form offers an additional interaction option; this is a design rationale, not evidence of superior psychological outcomes [14,15,16]. These features may shape user engagement [14,16]. Their value depends on the users, the design, and the setting [14].

Reviews of social robots in mental health suggest possible support for social connection and emotional wellbeing. However, many studies were small and mainly involved older adults with dementia [17,18]. Reviews of robots in physical rehabilitation have mainly examined movement, motor recovery, and physical function [19]. Design research also emphasizes matching interaction to users’ abilities and preferences, including alternative communication modes [16,18]. A recent home-use study included four adults with severe physical or neurological disabilities, whose interactions with a socially assistive robot involved emotional support and social participation [20]. Further work is needed to identify design requirements for combining mood check-ins, companionship, and mindfulness guidance at home for community-dwelling people with long-term physical disabilities.

Previous social robot design and codesign studies have mainly involved children or older adults. They have often focused on appearance or continued engagement [21,22,23,24]. These studies offer general design lessons. Further formative work is needed to understand how needs and preferences relevant to community-dwelling people with long-term physical disabilities could inform the physical form, dialogue, use setting, accessibility, and ethical features of a psychological support social pet robot.

We therefore conducted a formative user-informed design study to explore preliminary needs and preferences for a psychological support social pet robot intended for community-dwelling people with long-term physical disabilities. The clinical range represented in this initial sample was limited, and wider transferability was not tested.

### Objective

To address this aim, we conducted a formative user-informed design study with qualitative target user interviews. The aim was to identify preliminary needs and preferences and translate them into candidate requirements for a psychological support social pet robot for community-dwelling people with long-term physical disabilities. Its proposed role included brief mood check-ins, optional companion interaction, and mindfulness guidance at home. The robot was not intended to replace professional mental health or specialist medical care. A physical prototype was then built to show how the findings could inform a tangible design. The prototype was not evaluated in this study.

## Methods

### Study Design

We conducted a 3-phase formative user-informed design study with qualitative target user interviews. Phase 1 involved initial concept development by the authors and an external nursing advisor. Phase 2 involved interviews with 6 target users. Phase 3 involved an author-led design synthesis workshop. Four authors completed the live workshop using modified bilingual materials. They incorporated the interview themes and separate responses from one additional author and one target user participant. They retained responsibility for the final design decisions. These inputs formed an initial prototype plan. Table 1 summarizes the 3 study phases.

**Table 1.** Study Phases.

|  | **1. Initial concept development** | **2. Target user interviews** | **3. Workshop** |
|---|---|---|---|
| **People involved** | 5 authors and 1 external nursing advisor | 6 community-dwelling adults with long-term physical disabilities and 5 authors | 4 authors in the live workshop, with telephone input from 1 other author and 1 interview participant |
| **Activities** | Developed an early psychological support dialogue with mood check-in and mindfulness elements and a soft companion robot concept | Reviewed the initial concept in 3 qualitative interviews | Combined the interview findings and telephone input, reviewed the interaction guidance, and developed a prototype plan |

### Initial Concept Development

The initial setup phase formed an early concept for a psychological support social pet robot that could provide mood check-ins, companionship, and mindfulness guidance at home. This phase did not aim to define the final product. It created an initial concept that could be reviewed and revised through target user feedback.

As part of the early concept, the team designed the conversation and physical form of the robot. The conversation began with a mood check-in and linked users’ current feelings with supportive interaction and guided mindfulness practice. The mindfulness content drew on common parts of structured mindfulness programs [25]. A mood rating before each session was included to help users express their emotional and physical state. The robot used a soft panda form informed by prior work on companion robots with animal forms [26]. The panda form, soft exterior, and portable belt were early design ideas to be reviewed with target users.

### Target User Interviews

#### Participants and recruitment

We recruited 6 community-dwelling adults with long-term physical disabilities in Hong Kong. All 6 had inflammatory arthritis. They provided feedback on the initial robot concept from Phase 1. Eligible participants had ankylosing spondylitis, rheumatoid arthritis, or psoriatic arthritis. We excluded people with severe psychiatric illness. The team distributed a recruitment poster online. A study research assistant contacted potential participants through WhatsApp. Participants needed to be physically able to attend an interview in a campus meeting room. The authors confirmed eligibility before arranging the interviews. The sample comprised the 6 eligible people who were willing to participate during the recruitment period, and all completed the interviews. Disease severity, functional limitation scales, and digital technology experience were not systematically collected. This sample provided early feedback on the concept. The interviews consisted of 2 small group sessions with 3 and 2 participants and 1 individual session with 1 participant. Table 2 shows their characteristics. Five participants were women. This was 83.3% of the sample. The participants were aged 41 to 70 years. The mean age was 57.3 years. Two participants had rheumatoid arthritis. Two had ankylosing spondylitis. Two had psoriatic arthritis. The approximate time since diagnosis ranged from 2 to 24 years. This range was calculated from the reported diagnosis year and the interview year of 2026. As for socioeconomic status, 4 participants were not employed. This was 66.7% of the sample. Two participants worked part time. This was 33.3% of the sample.

**Table 2.**
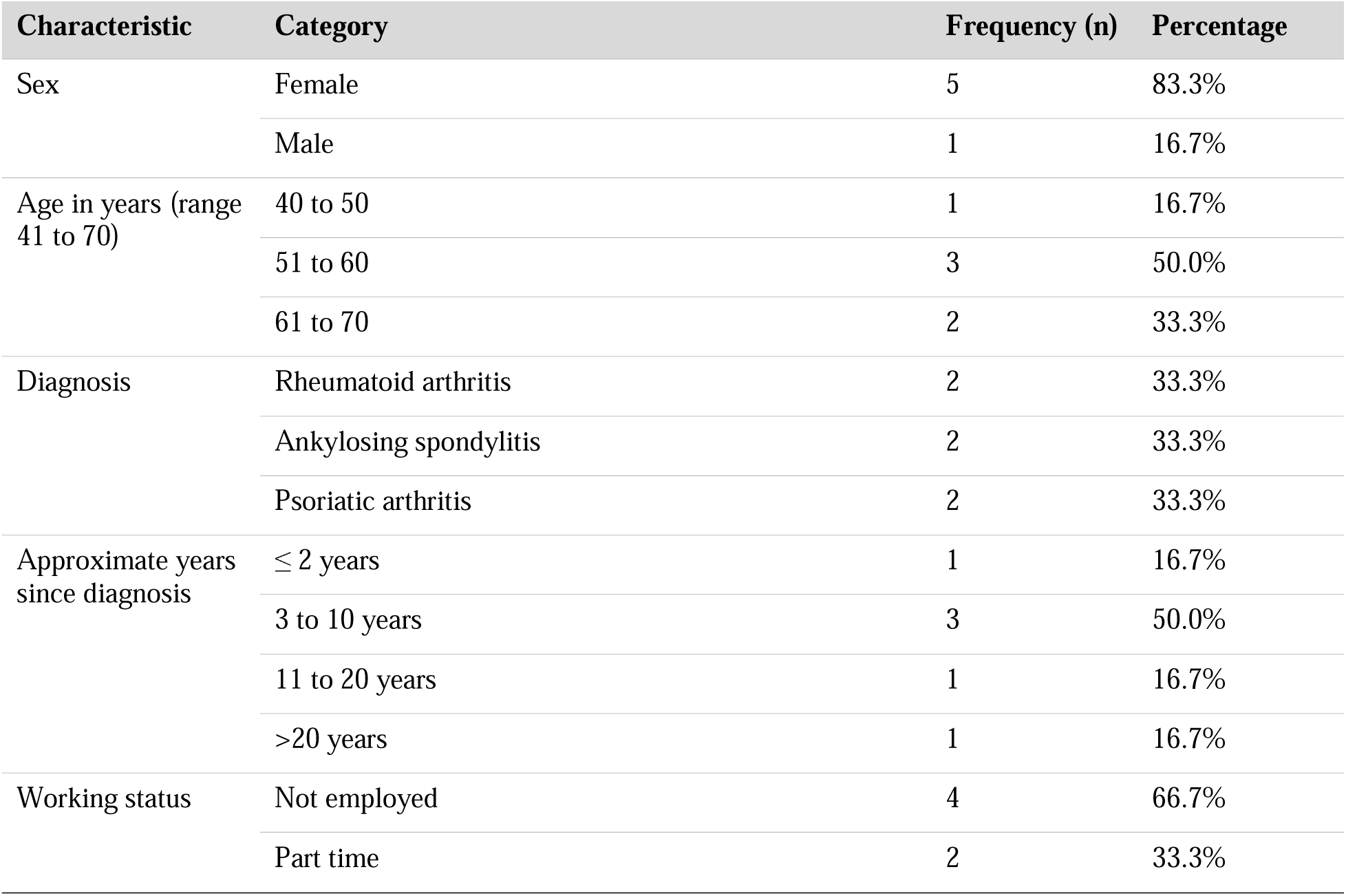
Participant Characteristics (N=6)

Five authors contributed to the design process. Their experience covered mental health, nursing, software, hardware, HCI, textile product design, product making, and physical design. An external nursing advisor supported the initial setup. This advisor was not an author or study participant.

#### Data collection

Three Cantonese interview sessions took place in Hong Kong in January 2026. The authors developed a semistructured interview guide for this study. The broad topics were informed by earlier participatory design research in which prospective users discussed their experiences with wellbeing practices and their views on the purpose, functions, and form of a robotic mental wellbeing coach [27]. The Social Robot Co-Design Canvases also informed questions about users, interaction, robot form, and context of use [28]. The authors adapted these topics to the study aim and the planned psychological support robot. During the interviews, participants viewed the planned conversation flow and early physical concept. The full questions and follow-up prompts are provided in Multimedia Appendix 2.

The session on January 23 lasted about 2 hours. It included 3 participants and 5 authors. The session on January 28 lasted 32 minutes and 4 seconds. It included 1 participant and 2 authors. The session on January 30 lasted 1 hour, 2 minutes, and 27 seconds. It included 2 participants and 2 authors. The study therefore used small group and individual interviews. One author led the interviews and made field notes. This author had no prior relationship with the participants. The participants gave permission for audio recording. A Hong Kong research assistant who was fluent in spoken Cantonese, written Chinese, and English transcribed the recordings word for word into Cantonese text. The original Cantonese transcripts were retained as the source data. Participants commented on the initial robot concept and identified features that could support or limit daily use. Their responses informed changes before the workshop.

#### Data analysis

The analysis followed a matrix-based qualitative content analysis approach [29,30]. Categories were developed inductively from the participants’ own wording within a deductive relevance-screening frame. The study’s design questions about function, form, and use context guided which material was treated as relevant. The original Cantonese transcripts were the source data for every analytic decision.

##### Segmentation

The stored matrix contained 536 utterance units initially classified as participant speech. An utterance unit was a continuous participant turn. Where a turn covered more than one distinct topic, it was divided into the smallest spans that each expressed a single idea about the concept, its functions, its form, or its use. Each unit received a stable identifier, such as M0113, so that later analytic steps and reported quotations could be traced back to their position in the transcript. Speech identified as interviewer material was read only to establish the context of a participant response.

##### Screening

The authors selected the screening keywords after reading the interview transcripts. A Python rule-based keyword screening program supported the initial relevance screen by matching the transcript text and associated researcher annotations. For a unit identified as participant speech, a match to any one keyword in either field produced a candidate-inclusion suggestion; a nonmatch was referred for manual judgment rather than automatically excluded. The coding author checked all 536 units against the original Cantonese transcripts and made the final inclusion decisions. The program did not determine the final categories or themes. The screening program processed the transcripts and annotations locally.

One author read all 536 units against the study aim. The author retained 205 units and excluded 331. Units were excluded when they carried no content about the needs, preferences, or use context of the proposed robot. Of the 331 excluded units, 17 (5.1%) were brief acknowledgment or filler responses with no independent meaning, such as a single word of agreement. In the stored suggestion field, 226 of the excluded units (68.3%) were marked as requiring manual judgment and were judged, on the coding author’s reading, not to address the study aim. A further 88 (26.6%) were marked as candidates for inclusion but were excluded on closer reading. A separate reason was not logged for each of these 331 units at this stage.

##### Contextual recheck

While charting the 205 retained units into the qualitative matrix, the coding author reread each unit a second time against its surrounding transcript context. This recheck removed 30 units (14.6% of the 205) that on closer reading did not support a reliable link to the robot concept, mindfulness practice, or the study’s design questions. Most of these were single clauses or fragments whose topic or referent could not be established from the surrounding text; a smaller number were a participant’s question addressed to another participant rather than a statement of that participant’s own experience or preference. These 30 units received no final category or theme and were not carried into the analysis below. The remaining 175 units informed the category and theme development that follows.

##### Category and theme development

The 175 units retained after the contextual recheck were charted into a qualitative matrix. Each row held one retained unit. The columns recorded the participant, the descriptive category, and the design question addressed. Categories were formed from the wording of the retained units. They were then compared across the full matrix and were merged, split, or renamed until each retained unit could be assigned without forcing. Related categories were grouped into subthemes, and the subthemes were grouped into 5 themes. Theme boundaries were checked by rereading the units in each category against the transcripts.

Participant identifiers were retained for traceability. Participant-level theme coverage is not reported because the recording equipment did not capture usable speech from one participant in the first group session, and the session formats gave participants unequal opportunities to speak.

##### Divergent cases

Less common and conflicting comments were retained when they were relevant to the study aim. They are reported with the themes in the Results and in Multimedia Appendix 1.

##### Verification of quotations and translations

Candidate Cantonese quotations were checked against the audio recordings, and this check is recorded in the qualitative evidence tracking file for the quotations used in Table 3 and in the text. The participant identifiers used in the reported quotations were also checked against the recordings. A bilingual author translated the checked quotations into English for use in Table 3 and in Multimedia Appendix 1.

**Table 3.** Interview Themes and Representative Quotations.

| Theme | Subtheme | Representative quotation (translated) |
|---|---|---|
| **Emotional support** | Family burden and concerns about expression | *“A friend may listen the first time, but if you say it 10 times…you do not want to bother them 10 times.” (P1, M0113)* |
|  | Companionship and social connection | *“Improving your mood really depends a lot on yourself. But it could provide companionship, so that you feel less lonely, right?” (P4, M0631)* |
| **Soft touch and appearance** | Softness and huggability | *“This one feels like cotton, but that one feels a little like jelly; it has some elasticity, and that feels better.” (P5, M0709)* |
|  | Cleaning and a replaceable outer layer | *“You cannot take it apart and wash it, can you? Actually, it would be enough if this piece could be pulled out.” (P4, M0422)* |
|  | Animal or companion-like form | *“Cats and dogs are the common ones; because I don’t particularly like pandas, and I don’t even have the slightest interest in visiting Ocean Park to see them.” (P2, M0197)* |
| **Mindfulness delivery** | User choice of mindfulness content | *“Of course, it is better to choose for yourself.” (P4, M0516)* |
|  | Practice timing and reminders | *“You have not done it today. Do you have 10 to 15 minutes to reward yourself or give yourself some space?” (describing a daily reminder; P2, M0340)* |
| **Context of use** | Public settings and privacy boundaries | *“I would not take this thing outside…because if you talked to it in the street, people would think I was crazy.” (P1, M0204)* |
|  | Daily life and symptom variation | *“We are in a lot of pain, many things in life can’t be done…I have to take medicine too, I have to rest too!” (P1, M0114)* |
| **Ease of use and accessibility** | Personalization and configuration | *“The images would appear automatically. That could be built into the robot; I really do not want to press buttons.” (P1, M0267)* |
|  | Voice-based volume adjustment | *“Could I tell it to speak louder, especially if controlling it myself is too difficult?” (P4, M0549)* |
|  | Weight and mobility | *“It cannot be so heavy. At my age, I cannot carry heavy things; if it is too heavy, the whole bag becomes heavy.” (P5, M0844)* |

*Note.* Subtheme labels are descriptive English renderings of the final categories in the qualitative matrix. A bilingual author translated the quotations from Cantonese. Ellipses indicate shortening for the table. Matrix IDs link each quotation to the qualitative evidence record. Fuller quotation context and divergent views are discussed below; Cantonese excerpts are provided in Multimedia Appendix 1.

##### Analytic responsibility

One author carried out the screening, the categorization, and the theme development, and made the final analytic decisions. This author also contributed to concept development, which may have influenced relevance judgments and interpretation. The study did not use a second independent coder and did not assess agreement between coders. The matrix and its unit identifiers were retained as an audit trail so that each reported theme and quotation could be traced to the source transcript. The implications of analysis by a single author are addressed in the Limitations.

### Workshop

#### Participants and inputs

Four authors attended the live workshop. Their backgrounds included nursing and mental health, software and human computer interaction, hardware development, and textile and physical product design. One additional author and one target user participant from the interview sample could not attend. Each answered the planned workshop questions by telephone before the live session. The target user did not join the live group discussion. The workshop inputs consisted of the interview themes, author notes, and the separate telephone responses.

#### Materials and procedure

The live workshop took place on March 17, 2026, and lasted about 1 hour. The workshop materials were adapted from Canvases 01 to 04 of the Social Robot Co-Design Canvases framework developed by Axelsson et al. [28]. The authors converted them into bilingual, study-specific materials. Canvas 01 addressed the problem space, including users, the design problem, and the use context. Canvas 02 addressed ethical considerations. Canvas 03 presented interaction guidelines. Its content had been determined before the workshop, so it was not open for new notes and was displayed only as a discussion prompt. Canvas 04 structured the prototype plan. The later canvases for environment, form, interaction, and behaviour were not used as separate stages.

During the workshop, the four attending authors reviewed the interview themes and telephone responses. They added relevant points to the materials that were open for note taking and discussed how the findings could inform the robot concept.

#### Data synthesis

The four attending authors synthesized the workshop notes into an initial prototype plan. The notes were analyzed separately from the interview transcripts and organized into robot display, interaction logic, and ethical considerations. An evidence map distinguished direct interview support, author interpretation, and technical decisions. The authors retained responsibility for the final design decisions. The workshop synthesis was not treated as target user consensus or as a separate validation exercise.

### Ethical Considerations

The study received ethics approval from the Institutional Review Board of The Hong Kong Polytechnic University (reference HSEARS20251103001; approval letter dated December 20, 2025). All participants gave written informed consent before data collection. Consent covered audio recording, English translation, publication of deidentified quotations, and use of deidentified transcript text in the study analysis. Each participant received a gift voucher worth HK$200. Speaker names were removed from the transcript text and annotations. These materials were processed and stored locally, with access limited to members of the research team.

## Results

### Initial Concept Development

The initial setup produced a portable psychological support companion concept for use in daily settings. It was intended to offer a mood check-in, companion-oriented interaction, and mindfulness practice chosen by the user. The early concept had a soft panda form, a carrying belt, and a simple spoken flow for checking mood and selecting mindfulness audio. Panel A of Figure 1 shows one representative early physical design. These features were starting points for the interviews. They were revised after participant feedback.

**Figure 1.**
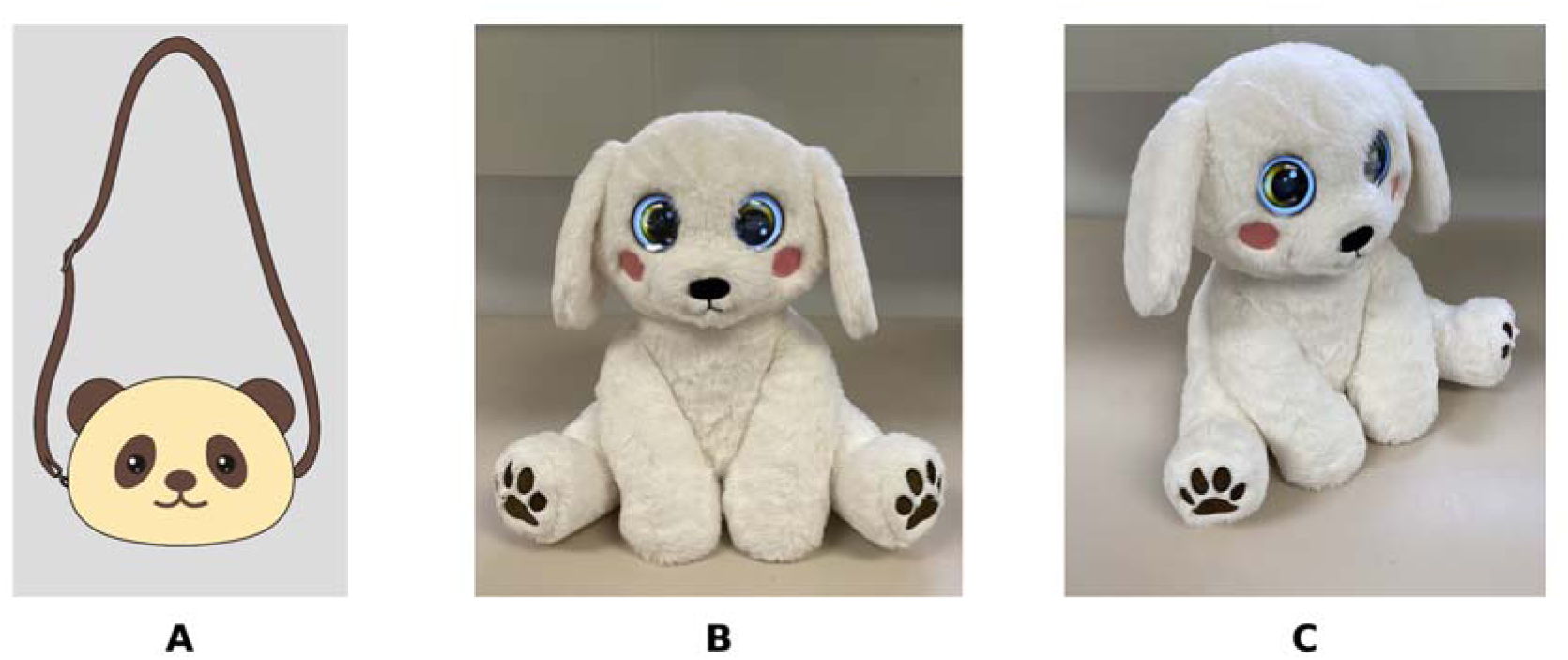
Design Evolution from Initial Concept to Prototype. *Note.* Panel A shows the initial portable panda concept with a carrying belt. Panels B and C show the final seated prototype in the form of a dog for use at home. The images illustrate author design decisions, not target user approval or prototype usability.

### Target User Interview Findings

Qualitative content analysis of the retained transcript units produced 5 themes. These were emotional support, soft touch and appearance, mindfulness delivery, context of use, and ease of use and accessibility. Table 3 presents the themes, their subthemes, and one representative quotation for each subtheme. The quotations are expanded in the text below. The research team used these findings, including divergent views, to prioritize home use and consider voice control as a candidate interaction option.

#### Emotional support

Participants described concerns about repeated emotional expression placing pressure on existing relationships and viewed companionship as a possible role for the robot. P1 explained, “A friend may listen the first time, but if you say it 10 times, even if they do not say they are annoyed, you do not want to bother them 10 times” (M0113). P4 limited the expected effect of a robot: “Improving your mood really depends a lot on yourself. But it could provide companionship, so that you feel less lonely, right?” (M0631). These comments supported an optional space for expression or companionship rather than a claim that the robot could improve mood or replace human support.

#### Soft touch and appearance

Participants discussed softness and huggability, cleaning, and animal or companion-like forms. When comparing materials, P5 said, “I think this feels very good. This one feels like cotton, but that one feels a little like jelly; it has some elasticity, and that feels better” (M0709). P4 focused on maintenance: “But with this one, you cannot take it apart and wash it, can you? Actually, it would be enough if this piece could be pulled out” (M0422). Preferences for animal form were not uniform. Referring to the initial panda concept, P2 said, “Cats and dogs are the common ones; because I don’t particularly like pandas, and I don’t even have the slightest interest in visiting Ocean Park to see them” (M0197). The theme therefore supported a soft and touchable companion form while leaving the animal type, visual style, and removable or washable exterior open for further testing.

#### Mindfulness delivery

Participants linked mindfulness delivery to practice format and content, timing, understandable guidance, and the boundaries of the system’s content. P1 questioned a narrow content range: “Then I might as well open YouTube and find some exercises. I think this thing could have more functions because it needs to give me information. If I suddenly have severe pain, or, like today, almost fall and need muscle exercises, why should I wait for it? Would it not be better if I could play a YouTube video and follow it?” (M0305). P2 explained that someone who was already very upset or frightened might not know what to do, even after being taught. P2 therefore proposed a regular daily reminder that could ask, “You have not done it today. Do you have 10 to 15 minutes to reward yourself or give yourself some space?” (M0340). These comments raised candidate requirements concerning content range, practice duration, and reminders. The authors limited the prototype to nonclinical psychological support; exercise information, instructor-led changes, and user-selected reminders were not implemented.

#### Context of use

Spoken interaction in public could attract unwanted attention, while pain, household work, medication, rest, and family schedules shaped the circumstances in which support could be used. P1 said, “Honestly, I would not take this thing outside, even if you made it into a box, because if you talked to it in the street, people would think I was crazy” (M0204). In a longer account of pain and household routines, P1 explained that “we are in a lot of pain, many things in life can’t be done” and later emphasized, “I have to take medicine too, I have to rest too!” (M0114). This account showed how symptoms and domestic responsibilities could constrain the timing and effort available for interaction. P4 also contrasted work and home schedules, whereas P5 said that carrying a doll while travelling or walking outside would be acceptable. These comments informed the authors’ decision to give priority to home and other private settings without treating home use as an exclusive requirement.

#### Ease of use and accessibility

Participants connected ease of use with personalization and configuration, voice-based volume adjustment, and weight and mobility. P1 described a device that could automatically show positive images when soothing music was selected: “When I play the music, it could automatically show happy images without my having to set them at that moment. I would know when I wanted the soothing music, and the images would appear automatically. That could be built into the robot; I really do not want to press buttons” (M0267). P4 asked, “It was at the maximum volume just now. Could I tell it to speak louder, especially if controlling it myself is too difficult?” (M0549). P5 emphasized physical burden: “It cannot be so heavy. At my age, I cannot carry heavy things; if it is too heavy, the whole bag becomes heavy” (M0844). These comments suggested configurable content and simple voice interaction as possible ways to reduce manual burden, together with attention to volume, weight, and handling. However, voice control may not provide enough access for every user or setting. Accessibility of the physical button, charging, weight, cleaning, fabric hygiene, volume, interruption handling, and nonvoice options requires user testing. The prototype therefore shows selected design responses. It is not a confirmed accessible system.

Interview input informed candidate design choices: P1 (M0204) informed private-use prioritization while P5 accepted outdoor use; P4 (M0549) suggested voice-based volume adjustment, distinct from P1’s automatic configuration request (M0267); and P4 (M0422) raised washability, which was not implemented. Softness predated the interviews; plain language and warm dialogue were author proposals. The form of a seated dog and the eye displays were later author or technical decisions. A locally deployed open-ended language model module was explored later during engineering development. It was not a direct target user requirement and was excluded from participant-facing use.

### Workshop

The workshop combined interview themes, author notes, and separate telephone responses from people who could not attend. Four authors worked with the modified workshop materials and produced an initial prototype plan. One additional author and one target user participant answered the workshop questions separately. The target user did not join the live group discussion. The workshop notes were synthesized under 3 design dimensions: robot display, interaction logic, and ethical considerations. Table 4 summarizes the main workshop outputs, related design decisions, and prototype implementation. The workshop was the live author synthesis stage following user-informed interviews and separate telephone input. It used target user findings and design input from different fields. The authors retained responsibility for the final design decisions.

**Table 4.** Workshop Outputs and Prototype Implementation.

| Workshop dimension | Main workshop output | Design decision and implementation status |
|---|---|---|
| **Robot display** | A physical companion suitable for use at home rather than a screen-only or wearable design. | A plush form representing a seated dog with animated eye displays was selected, and the carrying belt was removed. |
| **Interaction logic** | Warm and simple dialogue, clear mindfulness choices, and support for pauses or interruptions. | The prototype used a fixed dialogue flow, selectable mindfulness audio, and voice-based volume control. An interrupted dialogue could continue from its previous state, but audio playback could not be paused or resumed. Nonvoice alternatives for content selection and volume adjustment were not implemented. |
| **Ethical considerations** | Clear information about the robot's purpose, session length, data use, technical limits, and boundaries with professional care. | A spoken introduction to basic functions was included. Information about data use, technical limits, and professional boundaries was not fully implemented. |

For interaction logic, the workshop stated that the robot should guide mindfulness through warm and simple dialogue. Plain language was preferred over unexplained terms such as body scanning and mindfulness modules. The authors suggested repetition of user words and pauses for reflection. These features aimed to create a supportive rhythm. Their acceptability and effect have not been evaluated. The proof of concept used a fixed dialogue process with a small number of conditional paths. Users could select a mindfulness audio category. The robot could continue a dialogue from its previous state after an interruption. It could not pause or resume audio playback. The prototype did not include reminders selected by users or subsequent changes to content made by an instructor.

For ethical considerations, the workshop stressed clear information and realistic descriptions of system capabilities. The authors proposed that the robot should explain its purpose, session length, data practices, technical limits, and any optional function supported by an instructor. The prototype gave a spoken introduction to basic functions at first use. It did not provide information about data practices. The workshop also proposed a printed user manual and a reward system for progress. These ideas were not included in the prototype.

### Translation of Findings Into a Prototype

After the workshop, the authors built the physical proof of concept as a design outcome. Panels B and C of Figure 1 show the prototype. The early wearable panda concept became a seated companion in the form of a dog for use at home. The prototype was about 30 cm high. Its outer cover used plush fabric with a short pile, and its interior had soft filling. These materials gave it the feel of a plush toy. It also had animated eye displays, an internal rechargeable power supply, USB Type C charging, a physical power control, and voice interaction with volume adjustment. It gave a spoken introduction to basic functions at first use. Its interaction flow included a mood check-in, companion-oriented prompts, and user selection of mindfulness audio categories. The system then played an audio segment at random from the selected category. After an interruption, the robot could continue the dialogue from its previous state. Content selection and volume adjustment were available only through voice interaction; nonvoice alternatives were not implemented. Audio playback could not be paused or resumed. The complete set of 7 initial concepts and a rear view of the final prototype are provided in Multimedia Appendix 3. The prototype implemented only selected candidate requirements and also included author and technical choices not directly derived from participant data.

For transparency, a locally deployed Qwen language model was explored as a possible open-ended question module during engineering development. This module was not derived from the target user findings. Because predeployment testing of answer accuracy, harmful outputs, crisis responses, and privacy risks had not been completed, it was disabled for participant-facing use and was not treated as a design requirement or evaluated feature. Study participants did not use the module, and it processed no participant questions, audio, transcripts, or study data. Any future implementation would require separate safety, privacy, and usability evaluation.

## Discussion

### Principal Findings

#### From user input to design decisions

Target user interviews informed several parts of the early concept. The authors prioritized home and other private settings while retaining outdoor use as a possible preference. The soft exterior was part of the initial concept; plain language and dialogue details were developed in the authors’ workshop, and P4 suggested voice-based volume adjustment. The workshop then translated these findings into decisions about the physical form, dialogue, accessibility, and limits of the robot. Further development should involve target users in generating, selecting, and evaluating alternatives, as illustrated in participatory design research [21,23,24,28]. The findings raise questions for future research on possible roles for community nurses. Nurses could share patients’ home routines, support needs, and barriers. They could also help patients take part in later design and community service planning [11,12,31].

#### Emotional support and conversational interaction

Participants described concern that repeated emotional expression could burden relatives or friends. They considered the robot a possible source of companionship or a space for expression, but they did not assume that it would improve mood. We interpret this finding as support for optional, user controlled, and nonjudgmental interaction rather than for replacing human relationships or mental health care. Prior social robot research has similarly examined social connection, engagement, and mental wellbeing, but much of that evidence concerns older adults, including people with dementia, or general wellbeing populations rather than the present clinical sample [17,18,27]. An advantage of the prior literature is that it includes studies of actual robot interactions and wellbeing interventions, whereas the present study identified formative design requirements and did not test emotional outcomes. These findings offer formative evidence about preferences for companionship and optional interaction. In future community nursing practice, the robot could be offered as an optional home resource within patient education and self-management support. Community nurses could help users decide whether it suits their preferences, identify distress that needs human assessment, and maintain links to specialist medical and mental health services [7,11].

#### Soft embodiment, touch, and appearance

Participants valued a soft and touchable form, but they also raised cleaning, removable covers, and variation in appearance. We interpret softness as a feature that may support comfort, while appearance should remain adaptable rather than being treated as a universal preference. Prior work has shown the value of physical form and social presence and has examined a broad range of companion and home care robots [16,22,26]. However, many previous designs were developed for rehabilitation or older adult care, whereas the present concept was a psychological support social pet robot intended for community-dwelling people with long-term physical disabilities. These preferences concern companion robot design generally; their applicability to a wider range of community-dwelling people with long-term physical disabilities requires testing. In community care, nurses could contribute practical knowledge about comfort, cleaning, infection prevention, handling, and possible stigma before a device is introduced into the home. This possible role is consistent with nursing support based on individual needs and patient education tailored to the individual [11,12].

#### Home use, privacy, and daily routines

The interviews challenged the initial assumption that portability would be desirable. Participants described use at home or in another private setting, and their preferred timing and session length varied with work, household routines, pain, and privacy. We interpret this as a requirement to fit the intervention to daily life rather than asking users to adapt their routines to the robot. Previous research has the advantage of examining home robots through broad deployment frameworks and, in some studies, sustained codesign with older adults [21,22]. This work provides stronger insight into home use over the long term than the single interview phase of the present study. However, the populations and robots were different. The proposed robot was a stationary companion for brief psychological support, including mood check-ins, companion interaction, and mindfulness guidance, not a mobile home assistant with many functions. The contrasting views suggest that portability should remain a candidate design choice to be tested with users. In a future community nursing model, nurses could discuss home use as part of an assessment of symptoms, daily routines, caring responsibilities, privacy, and willingness to use spoken interaction.

#### Accessible interaction and physical effort

P1 discussed automatic content configuration, P4 requested voice-based volume adjustment, and P5 raised weight. P4 did not explicitly attribute control difficulty to arthritis, and P5 related the weight concern to age. We interpret these comments as showing that accessibility depends on the combined effort of controlling, hearing, moving, cleaning, and charging the robot. Voice control may reduce some manual demands, but it should not be the only option because speech, hearing, privacy, and environmental conditions differ. Rehabilitation and older adult robot research has similarly emphasized engagement, interaction, and the relationship between robot capability and the user’s functional context [16,18,22]. An advantage of that research is its inclusion of therapist knowledge. These comments identify access questions for further design of mental wellbeing support. A robot cannot provide useful mindfulness guidance if operating it becomes another burden or discourages use. Community nurses could help patients choose interaction methods that fit their preferences. In this way, community nursing could support access to home based mindfulness and emotional support [11,31].

#### Multiple psychological support functions and limits of the robot

The robot was designed as a psychological support companion rather than a mindfulness-only device. Mood check-ins, companionship, and mindfulness guidance served complementary roles. Participants discussed mindfulness format and timing, while the authors proposed plain and warm dialogue. Some participants also wanted exercise or health information. These requests raise an important design issue. More functions may make the robot seem more useful, but they may also make its role unclear. Existing mindfulness programs use structured content and support from trained facilitators [9,10,25]. Previous wellbeing robot studies have also stressed the need for clear design and ethical guidance [27,32]. The authors scoped the prototype as nonclinical psychological support rather than exercise or clinical advice. The robot should clearly state that it does not provide diagnosis, exercise advice, treatment, or crisis care. In future community nursing practice, nurses could review the support content, explain these limits, and direct users to professional care when needed [11,12].

### Limitations

Several limitations should be considered. First, the sample included only 6 target users. Most were women. They volunteered in response to a study invitation in Hong Kong. The sample included 3 types of inflammatory arthritis. The findings should not be generalized to all people with inflammatory arthritis or to the wider population of community-dwelling people with long-term physical disabilities, including those with different diagnoses, severity, access needs, causes of disability, or cultural contexts. Disease severity, functional limitation scales, and digital technology experience were not systematically collected, limiting interpretation of differences in design preferences. Second, the first of the study’s three interview sessions included 3 participants and 5 authors. This may have affected what participants felt able to say. The recording equipment did not capture usable speech from one participant in this session. The analysable material from the first interview session therefore came from only 2 of its 3 participants. Third, the user-informed design process relied on interviews and was partly asynchronous. Only one target user answered the workshop questions directly. Target users did not join the final group discussion or evaluate the completed prototype. The authors retained responsibility for the final design decisions. Fourth, one author carried out the screening, the categorization, and the theme development. The study did not use a second independent coder and did not assess agreement between coders. The author who led the analysis also contributed to the development of the concept under review, which may have influenced which material was judged relevant to the study aim. Individual exclusion reasons were not fully logged. The themes should therefore be viewed as a formative design synthesis. They are not a full or independently confirmed account of lived experience. Finally, the prototype only showed how the findings could inform a physical design. The prototype did not include a washable casing, user-selected reminders, controls to pause or resume audio, or full information about data use and technical limits. Nonvoice alternatives for content selection and volume adjustment were not implemented. The prototype was not evaluated at home, and its technical performance, privacy protection, and usability were not tested. An experimental local language model module was excluded from participant-facing use and was not evaluated. The study therefore provides no evidence about the safety or suitability of generative question answering and cannot confirm future feasibility or usability. Proposed nursing roles and implementation pathways were not evaluated.

### Conclusions

This formative study identified early needs, interaction preferences, and home-use considerations to inform a conversational psychological support social pet robot for community-dwelling people with long-term physical disabilities. The robot combined mood check-ins, optional companion interaction, and mindfulness guidance as complementary functions. User feedback and design synthesis by the authors changed the concept from a wearable companion to a device for use at home. Participant comments informed candidate preferences concerning touch, use settings, interaction effort, and timing; plain guidance, warm dialogue, and information about data use were author proposals. The physical prototype implemented only part of the candidate requirements alongside additional author or technical choices. It was not evaluated for usability, accessibility, acceptability, feasibility, or psychological benefit. Future research should involve community-dwelling people with a wider range of long-term physical disabilities. It should also examine usability, accessibility, privacy, and nursing value.

## Acknowledgments

The authors thank an external nursing advisor for support during initial concept development. The screening of transcript units, the development of categories and themes, the selection of quotations, the interpretation, and the final design implications were the work of the authors. A locally deployed Qwen language model was explored as a possible question module during engineering development but was excluded from participant-facing use. Generative artificial intelligence tools (Claude, Anthropic; Codex, OpenAI) assisted language editing and manuscript preparation across drafting and revision. The authors reviewed all AI-assisted text and take full responsibility for the content of the manuscript.

## Data Availability

The deidentified interview and workshop data supporting the findings of this study are available from the first author, Yufei Lu, upon reasonable request.

## Abbreviations

HCI: human computer interaction
JSON: JavaScript Object Notation

## Multimedia Appendix 1

*Qualitative Evidence Matrix and Mapping From Findings to Design*. Separate file.

## Multimedia Appendix 2

*Target User Interview Guide*. Separate file.

## Multimedia Appendix 3

*Initial Concept Gallery and Prototype Rear View*. Separate file.

