## Appendix 1 for "User-Informed Design of a Psychological Support Social Pet Robot for Community-Dwelling People With Long-Term Physical Disabilities: A Formative Study"

**Multimedia Appendix**

**Qualitative Evidence Matrix and Finding-to-Design Mapping**

This appendix summarizes the evidence base for the five interview-informed themes. Participant-level theme coverage is not reported because the recording equipment did not capture usable speech from one participant in the first group session, and the session formats gave participants unequal opportunities to speak. Each retained unit is counted once under the theme assigned to its final category in the summary matrix. These counts are not participant coverage or the total number of overlapping theme assignments.

| **Theme** | **Retained utterance units** | **Illustrative matrix IDs** | **Design interpretation and origin** | **Important limitation or divergent view** |
| --- | --- | --- | --- | --- |
| Emotional support | 32 | M0113; M0631 | P1 and P4 informed optional listening or companion interaction; this does not establish mood improvement or replacement of human or professional support. | P4 emphasized that mood improvement depends substantially on the individual. These comments concern possible companionship and do not establish psychological benefit. |
| Soft touch and appearance | 24 | M0197; M0422; M0709 | Softness was part of the initial concept. Participant feedback informed tactile preferences and interest in removable or washable outer components; washability was not implemented. | Preferences varied across animal form, colours, age-appropriateness, and portability; the dog-like form was not a universal preference. |
| Mindfulness delivery | 35 | M0516; M0340 | Participants discussed content choice, timing, and reminders. Plain-language guidance was an author proposal. User-selected reminders and instructor-supported adaptations were not implemented. | One participant questioned the value of a mindfulness-only system when immediate exercise information was wanted. Instructor-supported tailoring was raised by one participant only. |
| Context of use | 34 | M0114; M0204 | The authors prioritized home or other private settings while retaining outdoor use as a possible preference. Continuation of an interrupted dialogue state was implemented, but audio playback could not be paused or resumed; portability remains a candidate choice for future testing. | Some material conditionally supported carrying a companion outside; not all participants rejected portability. |
| Ease of use and accessibility | 50 | M0267; M0549; M0844 | P4 suggested voice-based volume adjustment; P1 suggested automatic content configuration. The prototype included voice-based content selection and volume adjustment and a physical power control. Nonvoice alternatives for content and volume were not implemented, and accessibility was not evaluated. | Voice interaction should not be the only possible control method for all users or environments. |

### Representative Verified Quotations

| **Theme** | **Subtheme (final category)** | **Participant and matrix ID** | **Cantonese quotation** | **English translation** |
| --- | --- | --- | --- | --- |
| Emotional support | Family burden and concerns about expression | P1, M0113 | 因為朋友呢你第一次講佢可能都會聽你講，當你講10次呢咁，雖然佢冇話好覺得厭煩，但係你都唔想去搞人10次啊。 | *“A friend may listen the first time, but if you say it 10 times, even if they do not say they are annoyed, you do not want to bother them 10 times.”* |
| Emotional support | Companionship and social connection | P4, M0631 | 改善啊，其實都好靠自己嘅改善情緒。不過佢可以咩嘅，即系陪伴好似冇咁孤單咁樣系嘛。 | *“Improving your mood really depends a lot on yourself. But it could provide companionship, so that you feel less lonely, right?”* |
| Soft touch and appearance | Softness and huggability | P5, M0709 | 一但係我覺得咁樣好好囉，因係佢嗰個感覺呢，你係咁落去呢？呢啲係好似有棉花嘅感覺嘛，但係佢感覺呢係一啲啫喱咁樣，即係點樣講，即係有個彈性，但得好啲囉。 | *“I think this feels very good. This one feels like cotton, but that one feels a little like jelly; it has some elasticity, and that feels better.”* |
| Soft touch and appearance | Cleaning and a replaceable outer layer | P4, M0422 | 但如果系呢只，但又唔會拆出嚟洗噶嘛系嘛？其實掹咗呢嚿嘢出嚟就得啦。 | *“But with this one, you cannot take it apart and wash it, can you? Actually, it would be enough if this piece could be pulled out.”* |
| Soft touch and appearance | Animal or companion-like form | P2, M0197 | 大路啲就系貓同狗咯，因為我冇特別中意熊貓，我連海洋公園去睇都冇呢個心嘅。 | *“Cats and dogs are the common ones; because I don’t particularly like pandas, and I don’t even have the slightest interest in visiting Ocean Park to see them.”* |
| Mindfulness delivery | Practice format and content | P1, M0305 | 咁我不如開個youtube攞啲練習出嚟做好過喇，即係呢嚿嘢我覺得可以有更多function，因為你要俾一啲資訊我呀嘛，令我突然間好痛呀，今日就突然之間就差啲跌倒喇，我係要做一啲肌肉練習嘅時候咁仲等你呀，我係咪自己撳個YouTube播條片出嚟咁咪跟住做，咁係咪仲理想呀？ | *“Then I might as well open YouTube and find some exercises. I think this thing could have more functions because it needs to give me information. If I suddenly have severe pain, or, like today, almost fall and need muscle exercises, why should I wait for it? Would it not be better if I could play a YouTube video and follow it?”* |
| Mindfulness delivery | User choice of mindfulness content | P4, M0516 | 咁梗係自己揀係好啲。 | *“Of course, it is better to choose for yourself.”* |
| Mindfulness delivery | Practice timing and reminders | P2, M0340 | 如果set咗15分鐘，其實亦都係好嘅即係，其實你心情好差，或者已經喺度驚緊呢，你教咗佢乜嘢佢都唔識做，但係如果佢定時每日15分鐘，咁佢就有個function remind你。今日冇做過喎，而家有冇10至15分鐘時間獎勵下你自己，或者等你自己有自己嘅空間。 | *“Setting it for 15 minutes would be good. When you are already very upset or frightened, you may not know what to do even after being taught. But if it reminds you at a regular time every day: ‘You have not done it today. Do you have 10 to 15 minutes to reward yourself or give yourself some space?’”* |
| Context of use | Public settings and privacy boundaries | P1, M0204 | 即係有冇apps呢個方向呢？因為真係老實講。我自己就唔會帶呢嚿嘢出街，even though你哋整一個咁嘅盒都唔會，因為你喺街你對住嚿嘢講嘅，人哋以為我痴線。 | *“Could there be an app option? Honestly, I would not take this thing outside, even if you made it into a box, because if you talked to it in the street, people would think I was crazy.”* |
| Context of use | Daily life and symptom variation | P1, M0114 | 我哋好痛，生活上好多嘢做唔到……大佬我都要食藥㗎，我都要唞㗎。 | *“We are in a lot of pain, many things in life can’t be done… Good grief, I have to take medicine too, I have to rest too!”* |
| Ease of use and accessibility | Personalization and configuration | P1, M0267 | 當我要播放音樂，咁就喺嗰個嘢度自己會彈翻出呢啲開心嗰啲……咁佢就自動彈啲相出嚟。咁就會用喺機械上咁就得啦，真係唔好撳掣囉。 | *“When I play the music, it could automatically show happy images without my having to set them at that moment… The images would appear automatically. That could be built into the robot; I really do not want to press buttons.”* |
| Ease of use and accessibility | Voice-based volume adjustment | P4, M0549 | 啱啱最大聲。咁可以叫佢大聲啲咁樣，佢自己控制太難嘅話呢？ | *“It was at the maximum volume just now. Could I tell it to speak louder, especially if controlling it myself is too difficult?”* |
| Ease of use and accessibility | Weight and mobility | P5, M0844 | 唔可以咁重，因為而家唔係年紀大呢，唔可以拎從嘢，咁如果你太重，呢整袋好重呢嗰隻就即係。 | *“It cannot be so heavy. At my age, I cannot carry heavy things; if it is too heavy, the whole bag becomes heavy.”* |

Subtheme labels are descriptive English renderings of the final categories in the qualitative matrix. The Cantonese quotations were checked against the recordings, and a bilingual author translated the verified quotations into English for use in the manuscript. Ellipses mark shortening of longer verified quotations.

### Translation Into the Proof-of-Concept Prototype

The study did not evaluate the prototype. The following table records features and candidate requirements with their implementation status; these include participant-informed ideas and author or technical decisions.

| **Design feature or candidate requirement** | **Status in the proof-of-concept prototype** |
| --- | --- |
| Soft, plush, and resilient exterior | Included; softness was part of the initial concept. |
| Eye displays capable of animations | Included as a prototype implementation decision. |
| Voice interaction and voice volume adjustment | Included for voice-based content selection and volume adjustment; no nonvoice alternatives for these functions were implemented. Accessibility was not evaluated. |
| Single physical power control | Included; accessibility was not evaluated. |
| User-selectable mindfulness-audio categories with random playback inside a selected category | Included; category choice reflected participant input, while random playback within a category was a technical implementation choice. |
| Continuation from an interrupted dialogue state | Included for dialogue state; audio playback could not be paused or resumed. Interruption handling was not evaluated with users. |
| First-use spoken introduction to basic functions | Included as an author/workshop proposal. |
| Removable or washable casing | Not included. |
| Carrying belt | Not included. |
| User-configurable reminders or session timing | Not included. |
| Instructor-supported adaptation of later content or plans | Not included. |
| Broader exercise or health-information functions, health monitoring, emergency contact, or progress-reward system | Not included. |
| Data-practice disclosure | Not included; proposed by the authors in the workshop. |

The detailed working records, verified Cantonese quotations, English translations, and negative cases are maintained in the project evidence workbook. Any public release of quotation-level or audit materials should follow the final ethics-approved data-access procedure.
