## Appendix 2 for "User-Informed Design of a Psychological Support Social Pet Robot for Community-Dwelling People With Long-Term Physical Disabilities: A Formative Study"

**Multimedia Appendix**

**Target User Interview Guide**

This file presents the content of the interview script. Formatting and minor wording were standardized for submission. The order and intended meaning of the questions and prompts were retained.

### Overall Aim

To understand what target users expected from a conversational robot intended to support emotional wellbeing, with particular attention to its physical display and audio interaction design.

### Part 1. Functions and Audio Interaction

The planned conversation flow and early box prototype were shown to participants.

#### Emotional Support Needs

**1.** What emotional concerns have you experienced that were caused by or related to inflammatory arthritis?

*Prompts:* anxiety, depression, loneliness, and quality of life.

**2.** Have you previously used audio content to manage emotional concerns? What types of audio content could help you manage these concerns?

*Prompts:* conversation, natural human speech, traditional poetry performance, music, and nature sounds.

*Follow up question:* In what situations would you prefer to use the audio method you mentioned, and why?

#### Mindfulness

**3.** How familiar are you with mindfulness?

If the participant was familiar with mindfulness: What type of mindfulness content would you prefer, and why?

If the participant was unfamiliar with mindfulness, the interviewer briefly explained that mindfulness involves intentionally paying attention to the present moment. Examples included mindful breathing, mindful eating, and a mindfulness body scan. The participant was then asked which type they would prefer and why.

**4.** How do you think mindfulness audio content could influence the emotional concerns you mentioned?

*Follow up prompts* explored why participants expected a positive influence, a negative influence, or no influence.

#### Functions and Interaction

**5.** What audio features would you expect from a robot designed to help with emotional concerns?

*Prompts:* guided conversation, self directed audio interaction, voice tone, session length, and frequency.

*Follow up questions included:*

- How would you feel about combining structured guidance with options for self directed conversation?

- Should sessions be freely selected or become available according to a schedule?

- What voice tone or speaking style would support you, and why?

- How long and how often should sessions take place?

- What speaking volume would be suitable when discussing personal emotional concerns?

The interviewer then explained that the proposed robot could guide users through prerecorded mindfulness and relaxation practices through conversation.

**6.** Considering the questions about audio interaction, what suggestions do you have for the current design?

*Prompts:* emotional concerns, mindfulness, and the audio interaction process.

**7.** Besides audio interaction, what other interaction channels would you expect in a future design?

*Prompts:* a visual display and physical control buttons.

### Part 2. Physical Display and Context of Use

#### Appearance

**8.** Thinking about the emotional concerns you mentioned, how could the appearance or design of a robot help address them?

*Follow up questions* asked about the size and colour of the robot.

#### Context of Use

**9.** In what situations would you use this robot?

*Prompts:* use at home during leisure, sleep preparation, or conversation, as well as use outside the home.

*Follow up question:* How do these situations relate to the emotional concerns you mentioned, and would the robot provide support or companionship?

**10.** How could this robot influence how your family or friends perceive you? How could this influence relate to the emotional concerns you mentioned?

*Follow up prompts* explored whether the robot could strengthen, weaken, or have no influence on relationships with family members or friends. Participants were asked about possible advantages and concerns.

The interviewer then showed the early appearance concept. It was a portable panda shaped plush design with bright colours, soft fabric, removable flower parts, and a keychain form that could be carried or hung from another item.

**11.** Considering the questions about appearance and context of use, what suggestions do you have for the current design?
