## Appendix 3 for "User-Informed Design of a Psychological Support Social Pet Robot for Community-Dwelling People With Long-Term Physical Disabilities: A Formative Study"

**Multimedia Appendix**

**Initial Concept Gallery and Prototype Rear View**

This appendix shows the complete set of early physical concepts considered during Phase 1 (Initial Concept Development) and a rear view of the final proof-of-concept prototype described in the Results (Translation of Findings Into a Prototype).

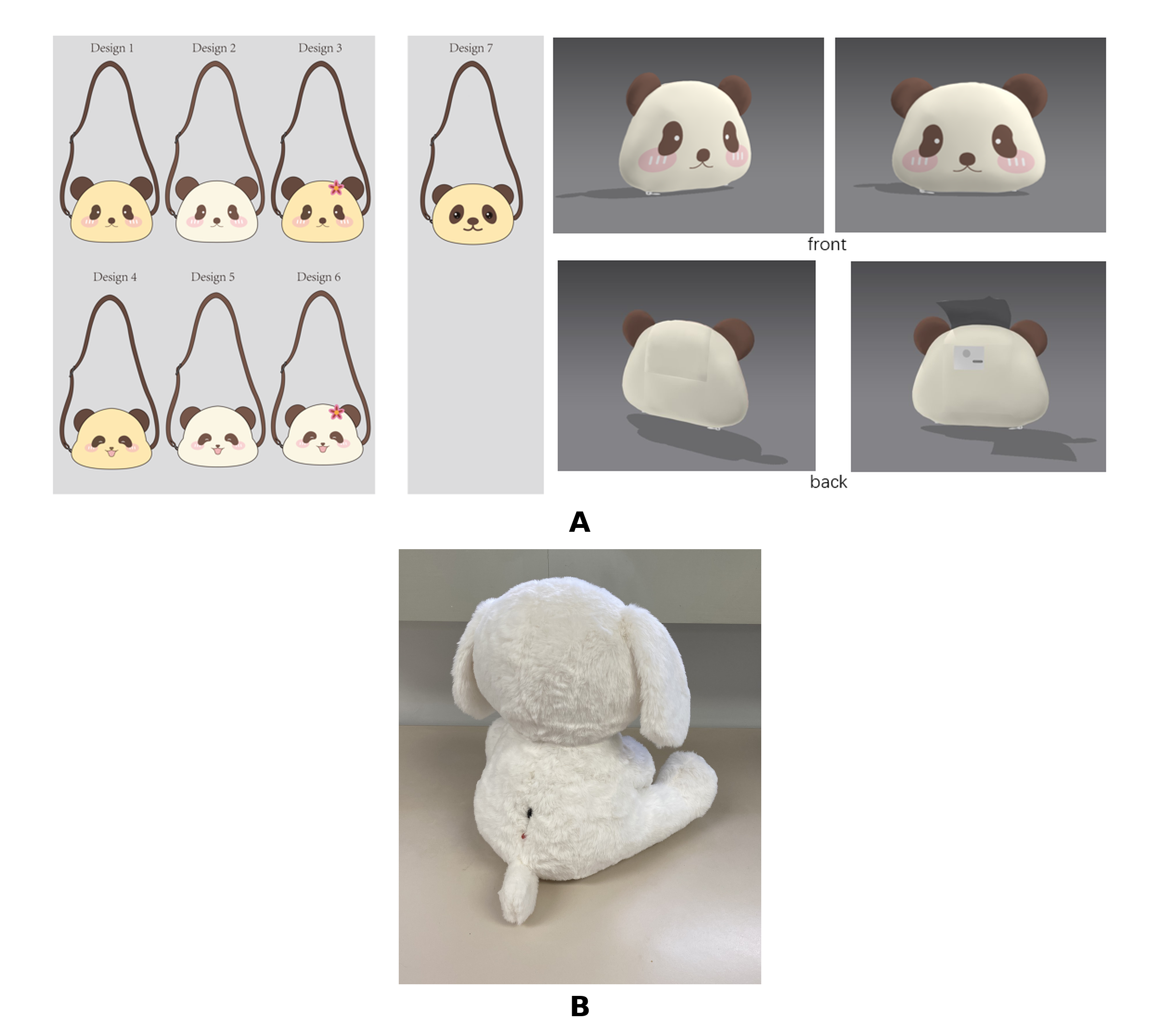

**Panel A.** The seven initial panda-form concepts (Design 1 to Design 7) developed during initial concept development, shown as flat design sketches. Design 7 is also shown as a 3-dimensional rendering from the front (2 views) and the back (2 views); this design informed the early physical concept used in the target user interviews.

**Panel B.** A rear view of the final seated dog-form prototype described in the Results, corresponding to the front and side views shown in Panels B and C of Figure 1 (Design Evolution From Initial Concept to Prototype).

*Note.* These images illustrate author design decisions during concept development and prototyping. They do not represent target user approval or an evaluation of prototype usability.
